# Development of the Tool for Advancing Practice Performance, a practice-level survey to assess primary care structures and processes

**DOI:** 10.1101/2024.11.28.24318176

**Authors:** Lorraine Kwok, Margaret M. Paul, Stephanie L. Albert, Daphna Harel, Saul B. Blecker, Bryan J. Weiner, Laura J. Damschroder, Deborah J. Cohen, Ann M. Nguyen, Donna R. Shelley, Carolyn A. Berry

**Author notes:** Corresponding author: (LK). LK and MMP are Joint First Authors.

## Abstract

Despite progress to define primary care practice transformation models, there remains gaps in translating evidence-based guidelines into routine clinical care. Primary care providers (MD, DO, NP, PA) and researchers need tools to assess modifiable factors that improve practice performance to inform practice transformation efforts. We aimed to develop a pragmatic tool for assessing practice-level primary care structures and processes that are associated with better care quality and clinical outcomes. We generated 314 candidate items for the Tool for Advancing Practice Performance (TAPP) using data from a comprehensive literature review, Delphi study, and qualitative interviews with high-performing practices. We used empirical criteria and expert review to eliminate redundancy and improve clarity via removing and retaining items. The retained items were formatted into a survey tool, and we further revised the tool based on feedback elicited from cognitive interviews and pilot testing with primary care providers and staff. The final candidate pool comprised 126 items after refinement and expert review. For the survey tool, we adapted and developed survey questions for each of the 126 items. Eight cognitive interview participants reviewed the tool and provided feedback on its content and language. Based on this feedback, we eliminated 13 items because they were poorly or incorrectly understood by participants, resulting in a 113-item tool. Fifteen participants pilot tested the tool and no additional items were eliminated. The TAPP is a novel, low-burden tool that researchers and primary care providers can use to identify areas for improvement at the practice-level. Practices and health systems could use the TAPP to assess their own performance and identify gaps in their structures and processes, and practice networks and health systems can use the tool to assess structures and processes at individual clinics, track this information over time, and evaluate its relationship to care quality and clinical outcomes.

## Introduction

Health care delivered according to evidence-based guidelines has been shown to lead to improvements in quality and clinical outcomes (1, 2). Over the past 15 years, new models of care such as the Chronic Care Model and the Patient-Centered Medical Home (PCMH) principles have been widely adopted throughout the US with the hope of transforming primary care delivery to improve care quality and outcomes (3, 4). Further, the articulation and dissemination of the PCMH principles by the American Academy of Family Physicians and other agencies sparked development of new tools designed for PCMH recognition and quality improvement processes, as well as the evaluation and accreditation of the level of “medical homeness” and its correlation with the triple aim: improving patients’ experience with care, improving population health, and reducing healthcare costs (5–7). There are a number of practice-level tools designed to measure overall primary care performance more generally, such as the Assessment of Chronic Illness Care (ACIC) (8) and the PCMHA (9), as well as tools that measure specific aspects of primary care believed to be predictive of performance including aspects such as teamwork (10, 11), organizational communication climate (12), and care coordination (13). There are several patient-reported measures used to assess the overall quality of primary care (14–23); however, patient-reported measures are labor intensive due to the burden of extensive primary data collection and analysis and therefore may be impractical for providers to conduct.

The underlying assumption behind the development and use of these tools is that the degree of fidelity to the model or desired state leads to a better score, and that the scores on the tools are directly associated with improved outcomes related to the various aspects of the triple aim. However, a notable and significant gap exists: most of the tools used to assess such performance have been neither rigorously developed nor assessed for reliability and validity, and none have established predictive validity with respect to care quality or patient outcomes (6, 7, 24–29). Therefore, despite progress made to define broad practice transformation theories and models and to develop tools to assess practice-level characteristics, there remains a gap in the translation of evidence-based guidelines into routine practice in primary care settings (30). Providers and researchers need tools to assess modifiable factors that improve primary care performance and inform efforts to transform care (2, 31).

Our overarching study aims to develop and validate a low-burden, practice-level tool that primary care providers and staff can use to assess the factors associated with better care quality outcomes for their practices. Our approach to data collection to inform tool development was guided by the Donabedian framework for assessing health services and quality of care with a focus on structures, or the context in which care is delivered, and processes, the methods by which health care is delivered, as the main drivers of these outcomes (32). The aim of this paper is to describe our comprehensive and data-driven approach to developing the Tool for Advancing Practice Performance (TAPP), including our initial data collection of evidence for specific structures and processes and the subsequent survey development, and cognitive interviews and pilot testing to refine the instrument.

## Methods

As part of our larger study, we first generated candidate items for the tool from three sources: 1) a comprehensive literature review of US-based research studies linking practice-level primary care structures and processes to quality and clinical outcomes; 2) a Delphi study in which 29 experts in the fields of primary care, health care management, and health services research rated the items generated from the literature review in terms of importance to chronic disease management and preventative care and feasibility of implementation (33); and 3) qualitative interviews of providers and quality improvement specialists from a national sample of 20 high performing primary care practices (34, 35). We engaged in a concept mapping exercise with 36 primary care providers, health system leaders, health services researchers, and implementation scientists to develop an empirically-derived structure to organize the items (36). The resulting structure consisted of 8 domains related to primary care: 1) Address Social Factors and Encourage Patient Engagement, 2) Reduce Clinical Risk Factors, 3) Provide Enhanced Care, 4) Expand Access to Care, 5) Provide Ancillary Services, 6) Establish Care Team Processes and Workflows, 7) Use Clinical Information Systems, and 8) Use Data and Evidence.

Following item generation and concept mapping, the development of the TAPP proceeded in four phases: 1) selection and refinement of items generated by our prior research, 2) survey tool development, 3) cognitive interviews to further refine the tool, and 4) pilot testing (see Fig 1). Throughout the study, the team met regularly with an expert panel that included implementation science researchers, health services researchers, and health system stakeholders; this expert panel was involved in key decision making during each phase of the research described in this paper. The study was approved by the New York University (NYU) Grossman School of Medicine Institutional Review Board.

**Fig 1.**
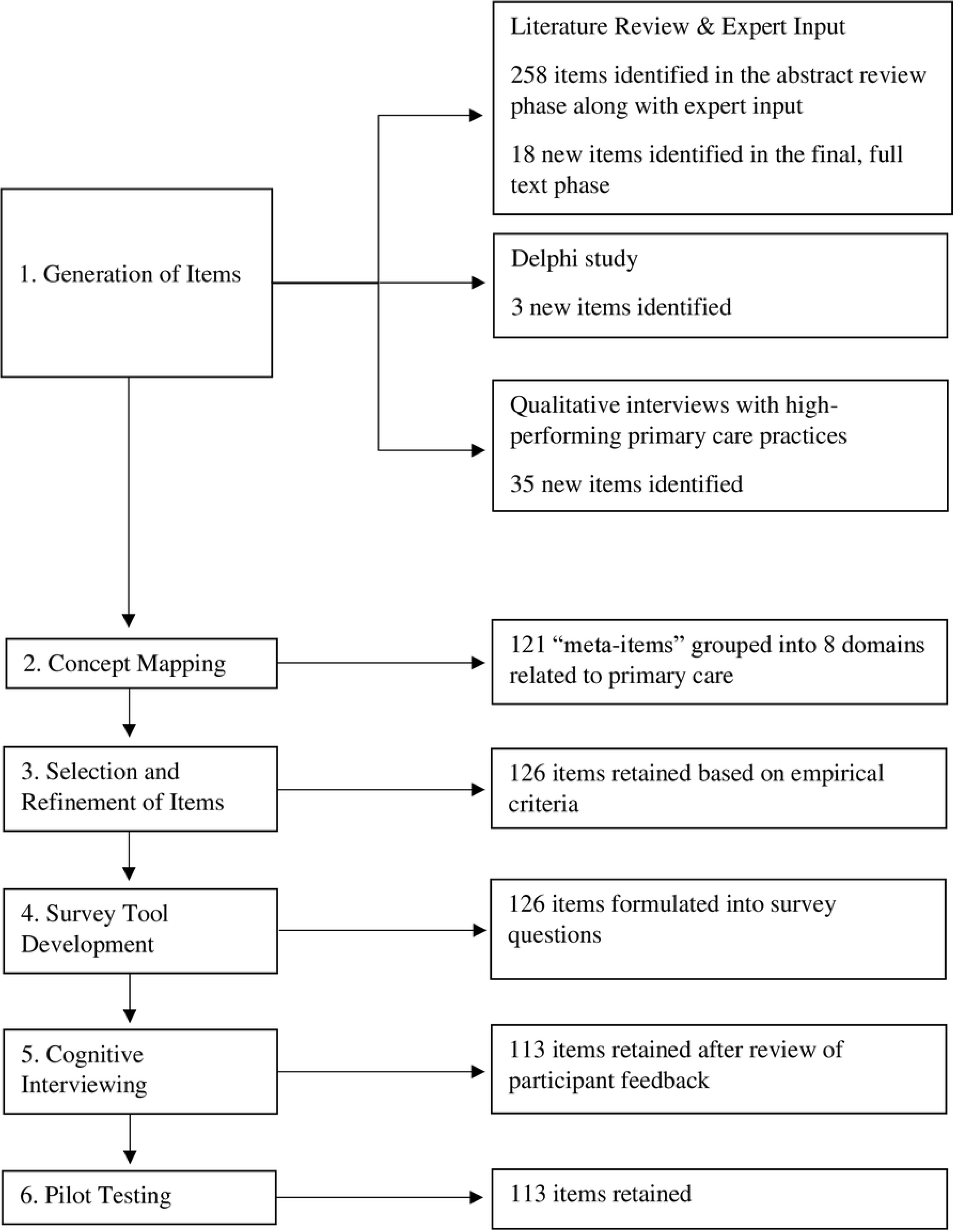
Flowchart of the development and content validation of the Tool for Advancing Practice Performance (TAPP).

### Generation of items

The primary purpose of the item generation stage was to empirically derive the content for our survey tool and to establish its content validity.

#### Literature review

In consultation with the Medical Library at NYU, we developed and refined our search strategy. We searched PubMed/MEDLINE, Embase, and Cochrane CENTRAL, and Web of Science for US-based research studies published in English between January 1, 2010 and the time the study began: December 31, 2018. We included any published study, commentary, or editorial that mentioned practice-level primary care structures and/or processes with the exception of articles published in languages other than English, studies based entirely outside the US, conference abstracts, and dissertations. We reviewed articles and collected data from our literature search in four stages: 1) title and abstract screening, 2) full-text article review, 3) full-text extraction, and 4) assessment of empirical evidence. Two members of the study team independently reviewed the content of each article at every stage and conflicts in opinion between pairs of reviewers were discussed as a group and resolved by consensus. During the title and abstract screening, the team screened articles and eliminated those that did not meet the inclusion criteria. In the second stage, the team reviewed the full text of articles to ensure retention of only those that met the inclusion criteria. We also reviewed the full text of articles in the review articles identified in the literature search and retained those that met the inclusion criteria.

#### Delphi study

Our Delphi study was designed to evaluate the impact and feasibility of primary care structures and processes identified through our literature review and input from our expert panel. We developed this study using the Conducting and Reporting Delphi Studies (CREDES) guidelines. The sample of participants included 29 primary care providers, health system leaders, and health services researchers in the U.S. In the first round of the study, participants rated the importance of each item with respect to both chronic disease management and screening and prevention. In the second round of data collection, participants reviewed aggregate results from the first round and were asked to come to consensus around the findings. Participants were also given the opportunity to contribute new items that were not included in our list of structures and processes, which led to the generation of 3 new items for consideration in our tool. Scores from the Delphi study were assigned to each structure and process in our list and used as evidence to retain or discard each item from the final survey set. The full Delphi study is further described in Albert et al (33).

#### Practice interviews

We conducted qualitative key informant interviews with providers and staff familiar with quality improvement activities. Our sample included 44 interviewees from 22 high performing practices located throughout the US. High performance was defined as meeting at least 2 of the following in 2018: among eligible patients, 70% prescribed aspirin use, 70% with controlled blood pressure (<140/90), 70% prescribed statin therapy, and 90% with controlled diabetes (HBA1c<9). We conducted semi-structured interviews, recorded our discussions, and coded the data to assess perspectives on structures and processes that contributed to their success. As with the Delphi scoring data, we incorporated data on the frequency with which structures and processes were discussed during these interviews as drivers of high performance on our outcomes of interest. This information contributed to our decision to retain or discard items for the tool.

### Selection and refinement of items

Data from three empirical data sources yielded 314 unique structures and processes, including 258 structures and processes identified from an initial broad review of abstracts in peer-reviewed journals and experience of the investigative team and expert panel and subsequently assessed in the Delphi study, and 56 additional items (18 from the final, full-text phase of the literature review, 3 suggested by experts from the Delphi study, 35 from the qualitative interviews). All 314 candidate items mapped onto eight domains related to primary care, which were derived from the concept mapping exercise that enabled development of an empirically-derived organizational structure for the 258 items included in the Delphi study.

We compiled all 314 items and the corresponding data in a matrix managed in Microsoft Excel. The reliance on empirical data from the literature review, Delphi study, and qualitative interviews rigorously established content validity of the tool. In order to refine the pool of structures and processes, we developed empirical criteria for removing and retaining items that relied on information from each mode of item generation. The guiding principle for inclusion in the TAPP was that each item had to have solid evidence, from at least one of our three sources of expert data, of a relationship to quality of care or patient outcomes.

Our Delphi process generated ratings on the perceived impact each item would have separately on chronic disease management or preventative care outcomes. We retained items if either the chronic disease management average or the screening and prevention average from the Delphi study was 3.5 out of 5 or above, an indication of group consensus on the item being of relatively high importance. We removed items if the Delphi score was below 3.5 for both chronic disease management and the screening and preventative means. Eliminated items could be reintroduced into the pool if four or more high performing practices mentioned them in the qualitative interviews or if there was at least one positive finding from the literature (i.e., the item was positively correlated with at least one relevant quality of care or clinical outcome variable).

We continued refining the set of items with regular meetings and input from our expert panel, until consensus was reached that all items were unique, clear, and specific. Items that were deemed redundant, lacked clarity, lacked specificity, or were considered universal or non-discriminatory were dropped from the list of candidate items.

### Survey tool development

We formatted a list of candidate items to create a low-burden survey tool (37), one that is parsimonious and user-friendly. Our goal in terms of burden was a tool that could be completed within 30 minutes and where items were unambiguous and easy to answer with accuracy. All but four of the survey items had a 3-point response set: yes, no, don’t know/unsure. Specific wording of questions was informed by an existing instrument (29) and developed de novo by the study team. We refined the tool through an iterative process with our expert panel and study team members to finalize formatting, organization, and phrasing of questions and response options. Our objective was to ensure that survey questions accurately reflected the structures and processes, and that each item was sufficiently brief and clear for primary care providers and staff.

### Cognitive interviews

We conducted eight cognitive interviews using videoconferencing from August to October 2021 to elicit feedback and to inform any further needed refinements. Participants included practice managers, office managers, providers, and quality improvement leaders from primary care practices that are part of two practice-based research networks in New York City and had previously participated in the qualitative interviews: New York City Department of Health and Mental Hygiene’s Bureau of Equitable Health Systems (BEHS), a bureau that supports New York City primary care practices in implementing patient-centered strategies for chronic disease prevention and management (38), and NYU Langone Health Faculty Group Practices, a network of three large multidisciplinary ambulatory care locations and more than 30 single and multi-specialty ambulatory care sites (39). We used concurrent think-aloud cognitive interviews where participants shared their thought process as they answered survey questions (37, 40). To facilitate this, participants shared the survey on their computer screens with the interviewers who followed along as the participants read each survey question aloud and narrated their thought processes in interpreting and answering each question. The interviewers probed for more explanation as needed. The use of a virtual meeting platform did not appear to compromise the quality of the feedback received. Interviews lasted for about one hour and participants received a $150 honorarium for their time and effort. We reviewed interview notes and through group consensus, determined survey revisions based on participants’ experiences.

### Pilot testing

Lastly, we piloted the final version of the TAPP with respondents from 15 practices to assess the mechanics of online survey administration, discern the time needed to complete the survey, clarify survey instructions, identify or confirm the roles most appropriate to engage in the survey, address any technical issues, and understand the overall experience taking the survey. The practices were recruited from three practice-based research networks: BEHS; DARTNet, a nonprofit organization that fosters research and supports collaboration with health care organizations, academic medical centers, and providers to extract data from a range of electronic health record (EHR) systems and use those data for quality improvement and research (41); and OCHIN, a nonprofit organization that provides an EHR system to over 500 federally qualitative health centers and community health centers nationwide (42). In addition to providing written feedback at the end of the survey, participants had the option to provide additional feedback via a scheduled call with study team members. Participants received a $150 honorarium for their time and effort.

## Results

### Selection and refinement of items

After applying criteria for removing and retaining items to the potential pool of items (n=314), 188 items remained in the candidate pool of items, which included 125 items with a Delphi score of 3.5 or higher for chronic disease management or screening and preventative care and 63 items that did not meet criteria in the Delphi process but were reintroduced because four or more practices mentioned the item in the qualitative interviews and/or there was at least one positive finding from the literature. Of the 63 items that were reintroduced, 43 items were mentioned by 4 or more practices in the qualitative interviews; 12 items had at least one positive finding from the literature; 5 items were mentioned in the qualitative interviews and had at least one positive finding from the literature; and 3 new items were suggested by experts in the Delphi study.

During the expert review of the 188 candidate items, we eliminated 70 items including 23 items due to redundancy; 8 for lack of clarity/specificity; 10 for being universal/non-discriminatory; 1 with low feasibility ranking from the Delphi study; 1 that reflected one of our outcomes of interest rather than a structure or process (i.e., documentation of patients’ smoking status); and 27 conceptually similar items were combined to form new item. This reduction process resulted in a refined pool of 118 items.

Further, we added eight items that were not previously included in our original list of structures and processes. Through expert review and consensus, we included five new items that were split off from existing items that were bundled (e.g., ‘Practice routinely screens patients for alcohol, drug, and tobacco use’ was split into three separate items to individually capture alcohol, drug, and tobacco use screening); two new items that capture additional staff that are part of a care team; and one new item that captures the use of a visual dashboard to manage preventative care for patients.

After distilling and refining the items, the final candidate pool comprised 126 items.

### Survey tool development

For the survey tool, we adapted and developed survey questions for each of the 126 items retained in our item pool. We improved the wording of the questions, response options, and organization of the tool based on input from our expert panel and meetings with study team members. To enhance the ease of completion, we used a 3-point response scale: yes, no, and don’t know/unsure. This scale requires less cognitive burden compared to estimates of degree or frequency of implementation. Questions were organized within the eight domains identified in our concept mapping exercise.

### Cognitive interviews

Our eight cognitive interview participants reviewed the 126-item tool and provided feedback on the content, item structure, and language as they reviewed each question. We reviewed and discussed the respondents’ feedback in real time and edited the survey as we progressed through the interviews. Ultimately, we eliminated 13 items from the tool because they were poorly or incorrectly understood by respondents in the interviews, which resulted in a tool with 113 items. Additionally, language for 23 items was modified for clarity based on feedback from the cognitive interviews (e.g., “In this practice, how often do patients see the same PCP?” was modified to “In this practice, how often do patients see the same PCP (other than vacation time)?).

### Pilot testing

Fifteen participants including executive directors, practice managers, office managers, providers, and quality improvement leaders piloted the 113-item tool. On average, the participants completed the TAPP survey in 30 minutes, meeting our threshold for low burden. The time it took to complete the TAPP ranged from 20 to 90 minutes. In addition to providing written feedback at the end of the pilot survey, two participants opted to provide additional feedback via a scheduled call with the study team. No additional items were eliminated beyond those eliminated based on feedback from the cognitive interviews. Survey instructions were modified slightly, and technical issues related to online survey administration were optimized for data collection.

## Discussion

The TAPP is the first primary care practice assessment tool based on empirical evidence of relationships between its items and clinical quality and outcome measures, thus establishing strong content validity. The tool assesses 113 discrete primary care structures and processes that contribute to high performance and can be completed within 30 minutes by one or multiple primary care practice staff including practice managers, office managers, providers, and quality improvement leaders. The TAPP was developed through multiple phases of research including selection and refinement of items generated from a comprehensive literature review, a Delphi study, and qualitative interviews with high-performing practices; development and refinement of survey items; cognitive interviewing; and pilot testing. Expert review of items from multiple data sources, cognitive interviewing, and pilot testing resulted in a refined set of primary care structures and processes that, if changed, could reasonably be expected to lead to improvements in health care quality and clinical outcomes related to chronic disease management and prevention. We know of no other measures of primary care practice performance that were developed with such systematic and comprehensive content validity. Moreover, the tool is relatively short and easy to complete with majority yes/no/unsure response set and thus represents a low burden on practices. The TAPP is free and publicly available for review and use in research and practice (see S1 File).

The development of the TAPP underscores the value of drawing upon multiple methods and data sources. While there was overlap between items identified in the literature, by Delphi experts, and providers and quality improvement specialists at high performing practices, we found that each source contributed key items that would have been underrepresented or absent if we had relied on a single source. Three new items emerged only through the Delphi panel study (e.g., the practice screens all patients using a standardized tool to identify past trauma; the practice has providers and staff trained in implicit bias), and 35 new items emerged from the qualitative interviews with practices (e.g., practice partners with onsite or local dental services; providers routinely update problem list in EHR). Additionally, during the Delphi exclusion stage, 37 items were “rescued” because they were mentioned by four or more practices in the qualitative interviews, suggesting that primary care experts and on-the-ground primary care providers and staff may have differing views on what items most affect care quality and chronic disease management. Efforts to develop and validate complex measurement tools in primary care should consider methods to gather data from multiple data sources to ensure comprehensive identification of primary care elements of interest.

The TAPP captures complex elements of primary care, such as structures and processes thought to be readily changeable at the practice level (i.e., without external support). The tool is a new resource for researchers and primary care providers to use for assessing care structures and processes and identifying areas for improvement at the practice-level. Practices could use the TAPP as a self-assessment of their performance and identify gaps in their structures and processes. Practice networks and health systems could use the TAPP to assess the structures and processes in place at individual clinics, track this information over time, and assess its relationship to quality and clinical outcomes. Additionally, the TAPP has the potential to measure the effectiveness of interventions designed to improve practice performance. Future work will evaluate the scoring and reliability of the TAPP based on data collected from practices across the US as well as validate the tool against care quality and patient outcomes. Further research is also needed to incorporate recent advancements in the use of telehealth in the delivery of routine primary care.

## Data Availability

All relevant data are within the manuscript.

## Acknowledgements

The authors thank the participants from the cognitive interviews and pilot testing for reviewing and providing feedback on the survey tool.

## Supporting information

S1 File. The Tool for Advancing Practice Performance (TAPP).

